# Effect of a one-month lockdown on the epidemic dynamics of COVID-19 in France

**DOI:** 10.1101/2020.04.21.20074054

**Authors:** Lionel Roques, Etienne Klein, Julien Papaïx, Antoine Sar, Samuel Soubeyrand

## Abstract

The COVID-19 epidemic started in the Hubei province in China in December 2019 and then spread around the world reaching the pandemic stage at the beginning of March 2020. Since then, several countries went into lockdown. We estimate the effect of the lockdown in France on the contact rate and the effective reproduction number *R*_*e*_ of the COVID-19. We obtain a reduction by a factor 7 (*R*_*e*_ = 0.47, 95%-CI: 0.45-0.50), compared to the estimates carried out in France at the early stage of the epidemic. We also estimate the fraction of the population that would be infected by the beginning of May, at the official date at which the lockdown should be relaxed. We find a fraction of 3.7% (95%-CI: 3.0-4.8%) of the total French population, without taking into account the number of recovered individuals before April 1st, which is not known. This proportion is seemingly too low to reach herd immunity. Thus, even if the lockdown strongly mitigated the first epidemic wave, keeping a low value of *R*_*e*_ is crucial to avoid an uncontrolled second wave (initiated with much more infectious cases than the first wave) and to hence avoid the saturation of hospital facilities. Our approach is based on the mechanistic-statistical formalism, which uses a probabilistic model to connect the data collection process and the latent epidemiological process, which is described by a SIR-type differential equation model.

## 1 Introduction

COVID-19 epidemic started in the Hubei province in China in December 2019 and then spread around the world reaching the pandemic stage at the beginning of March 2020 [1]. To slow down the epidemic, several countries went into lockdown with different levels of restrictions. In the Hubei province, where the lockdown has been set long before the other countries (on January 23), the epidemic has reached a plateau, with only sporadic new cases by April 15 (from the data of Johns Hopkins University Center for Systems Science and Engineering [2]). In France, the first cases of COVID-19 were detected on January 24, and the lockdown has been set on March 17.

The basic reproduction number *R*_0_ corresponds to the expected number of new cases generated by a single infectious case in a fully susceptible population [3]. Several studies, mostly based on Chinese data, aimed at estimating the *R*_0_ associated with the COVID-19 epidemic, leading to values from 1.4 to 6.49, with an average of 3.28 [4]. As the value of *R*_0_ can be interpreted as the product of the contact rate and of the duration of the infectious period, and since the objective of the lockdown and associated restriction strategies are precisely to decrease the contact rate, an important effect on the number *R*_*e*_ of secondary cases generated by an infectious individual is to be expected. This value *R*_*e*_ is often referred to as ‘effective reproduction number’, and corresponds to the counterpart of *R*_0_ in a population that is not fully susceptible [5]. If *R*_*e*_ *>* 1, the number of infectious cases in the population follows an increasing trend, and the larger *R*_*e*_, the faster this trend. On the contrary, if *R*_*e*_ *<* 1, the epidemic will gradually die out. The study [6] showed that containment policies in Hubei province indeed led to a subexponential growth in the number of cases, consistent with a decrease in the effective reproduction number *R*_*e*_.

Standard epidemiological models generally rely on SIR (Susceptible-Infected-Removed) systems of ordinary differential equations and their extensions (for examples of application to the COVID-19 epidemic, see [7, 8]). With these models, and more generally for most deterministic models based on differential equations, when the loss of observation due to the observation process is heavy, specific approaches have to be used to bridge the gap between the models and the data. One of these approaches is based on the mechanistic-statistical formalism, which uses a probabilistic model to connect the data collection process and the latent variable described by the ODE model. Milestone articles and textbook have been written about this approach or related approaches [9], which is becoming standard in ecology [10, 11]. The application of this approach to human epidemiological data is still rare.

In a previous study [12], we applied this framework to the data corresponding to the beginning of the epidemic in France (from February 29 to March 17), with a SIR model. Our primary objective was to assess the infection fatality ratio (IFR), defined as the number of deaths divided by the number of infected cases. As the number of people that have been infected is not known, this quantity cannot directly measured, even now (on April 15). The mechanistic-statistical framework allowed us to compute an IFR of 0.8% (95%-CI: 0.45-1.25%), which was consistent with previous findings in China (0.66%) and in the UK (0.9%) [13] and lower than the value previously computed on the Diamond Princess cruse ship data (1.3%) [14]. In this previous study, we also computed the *R*_0_ in France, and we found a value of 3.2 (95%-CI: 3.1-3.3). Although the number of tests at that stage was low, an advantage of working with the data from the beginning of the epidemic was that the initial state of the epidemic was known.

Here, we develop a new mechanistic-statistical approach, based on a SIRD model (*D* being the dead cases compartment), in the aim of

- estimating the effect of the lockdown in France on the contact rate and the effective reproduction number *R*_*e*_;
- estimating the number of infectious individuals and the fraction of the population that has been infected by the beginning of May (at the official date at which the lockdown should be relaxed).

## 2 Materials and Methods

### 2.1 Data

We obtained the number of positive cases and deaths in France, day by day from Santé Publique France [15], from March 31 to April 14. We obtained weekly data on the number of individuals tested (in private laboratories and hospitals) from the same source. We assumed that during each of these weeks the number of tests per day was constant. This assumption is consistent with the small variations between the number of tests during the first week (111 690) and the second week of observation (132 392). As the data on the number of positive cases are not fully reliable (fewer cases during weekends with a rebound on Monday), we smoothed the data with a moving average over 5 days. Official data on the number of deaths by COVID-19 since the beginning of the epidemic in France only take into account hospitalised people. About 728 000 people in France live in nursing homes (EHPAD, source: DREES [16]). The number of deaths in these structures has only been reported recently, and cannot be obtained day by day. Latest data from Santé Publique France indicate a total number of 10 643 deaths at hospital and 6 524 deaths in nursing homes by April 15. The total number of deaths therefore corresponds to about 1.6 times the number of deaths at hospital. The same factor had been estimated in [12] based on local dataset in the French Grand Est region.

### 2.2 Mechanistic-statistical framework

The mechanistic-statistical framework consists in the combination of a mechanistic model that describes the epidemiological process, a probabilistic observation model and an inference procedure.

#### 2.2.1 Mechanistic model

The dynamics of the epidemic are described by the following SIRD compartmental model:

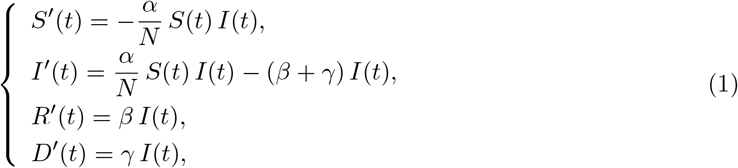

with *S* the susceptible population, *I* the infectious population, *R* the recovered population, *D* the number of deaths due to the epidemic and *N* the total population. For simplicity, we assume that *N* is constant, equal to the current French population, thereby neglecting the effect of the small variations of the population on the coefficient *α/N*. The parameter *α* is the contact rate (to be estimated) and 1*/β* is the mean time until an infectious becomes recovered. Based on the results in [17], the median period of viral shedding is 20 days, but the infectiousness tends to decay before the end of this period: the results in [18] show that infectiousness starts from 2.5 days before symptom onset and declines within 7 days of illness onset. Based on these observations we assume here that 1*/β* = 10 days. The parameter *γ* corresponds to the death rate of the infectious (to be estimated).

#### Initial conditions

The model is started at a date *t*_0_ corresponding to April 1st. The initial number of infectious *I*(*t*_0_) = *I*_0_ is not known and will be estimated. The total number of recovered at time *t*_0_ is also not known. However, as the compartment *R* has no feedback on the other compartments, we may assume without loss of generality that *R*(*t*_0_) = 0, thereby considering only the new recovered individuals, starting from the date *t*_0_. We fixed *D*(*t*_0_) = 3523, the number of deaths at hospital by March 31. The initial *S* population at the beginning of the period, should still be close to the total French population: by March 31 only 52 128 cases had been observed in France, corresponding to 0.08% of the total population. A factor 8 had been estimated in [12] between the cumulated number of observed cases and the actual number of cases at the beginning of the epidemic. Even though this factor may have changed, e.g. increased by a factor 5, this means that the proportion of the total population that has been infected by March 31 is still small. We may assume any value for *S*(*t*_0_) between 60 10^6^ and 67*·*10^6^ without changing much the results of our study (as *S/N* remains close to 1). For our computation, we assumed that *S*(*t*_0_) = 66*·*10^6^, corresponding to about 98.5% of the French population.

#### Numerical method

The ODE system (1) was solved thanks to a standard numerical algorithm, using Matlab^®^ *ode45* solver.

#### 2.2.2 Observation model

The number of cases tested positive on day *t*, denoted by 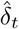, is modelled by independent binomial laws, conditionally on the number of tests *n*_*t*_ carried out on day *t*, and on *p*_*t*_ the probability of being tested positive in this sample:

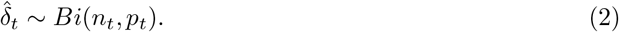

The tested population consists of a fraction of the infectious cases and a fraction of the susceptibles: *n*_*t*_ = *τ*_1_(*t*) *I*(*t*) + *τ*_2_(*t*) *S*(*t*). Thus,

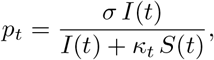

with *κ*_*t*_ := *τ*_2_(*t*)*/τ*_1_(*t*), the relative probability of undergoing a screening test for an individual of type *S vs* an individual of type *I*. We assumed that the ratio *κ* was independent of *t* over the observation period. The coefficient *σ* corresponds to the sensitivity of the test. In most cases, RT-PCR tests have been used and existing data indicate that the sensitivity of this test using pharyngeal and nasal swabs is about 63 − 72% [19]. We assumed here *σ* = 0.7 (70% sensitivity).

Each day, the number of new observed deaths (excluding nursing homes), denoted by 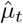, is modelled by independent Poisson distributions conditionally on the process *D*(*t*), with mean value *D*(*t*) − *D*(*t* − 1) (which measures the daily increment in the number of deaths):

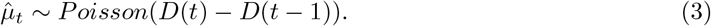

Note that the time *t* in (1) is a continuous variable, while the observations 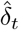 and 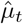 are reported at discrete times. For the sake of simplicity, we used the same notation *t* for the days in both the discrete and continuous cases. In the formulas (2) and (3) *I*(*t*), *S*(*t*) and *D*(*t*) are computed at the end of day *t*.

#### 2.2.3 Statistical inference

The unknown parameters are *α, γ, κ* and *I*_0_. We used a Bayesian method [20] to estimate the posterior distribution of these parameters.

#### Computation of the likelihood function

The likelihood ℒ is defined as the probability of the observations (here, the increments 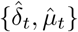) conditionally on the parameters. Using the observation models (2) and (3), and using the assumption that the increments 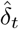 and 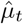 are independent conditionally on the underlying SIRD process and that the number of tests *n*_*t*_ is known, we get:

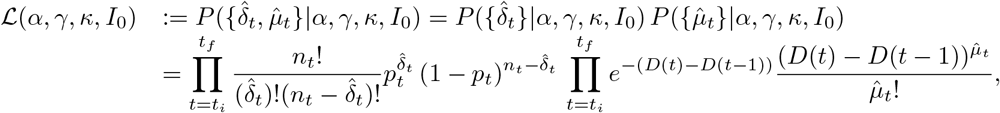

with *t*_*i*_ the date of the first observation and *t*_*f*_ the date of the last observation. In this expression ℒ(*α, γ, κ, I*_0_) depends on *α, γ, κ, I*_0_ through *p*_*t*_ and *D*(*t*).

#### Posterior distribution

The posterior distribution corresponds to the distribution of the parameters conditionally on the observations:

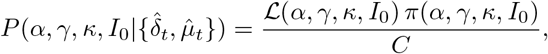

where *π*(*α, γ, κ, I*_0_) corresponds to the prior distribution of the parameters (detailed below) and *C* is a normalization constant independent of the parameters.

#### Prior distribution

Regarding the contact rate *α*, the initial number of infectious cases *I*_0_ and the probability *κ*, we used independent non-informative uniform prior distributions in the intervals *α* ∈ (0, 1), *I*_0_ ∈ (1, 10^7^) and *κ* ∈ (0, 1). To overcome identifiability issues, we used an informative prior distribution for *γ*. This distribution, say *f*_*g*_, was obtained in [12] during the early stage of the epidemic (*f*_*g*_ is depicted in the Appendix, Fig. S1). In [12], the number of infectious cases *I*_0_ at the beginning of the epidemic was known (equal to 1), and did not need to be estimated. Thus, we estimated in [12] the distribution of the parameter *γ* by computing the distribution of the infectious class and using the formula *D*^*′*^(*t*) = *γ I*(*t*) together with mortality data (which were not used for the estimation of the other parameters, unlike in the present study). Finally, the prior distribution is defined as follows:

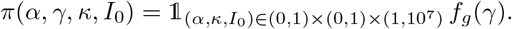

The numerical computation of the posterior distribution is performed with a Metropolis-Hastings (MCMC) algorithm, using 5 independent chains, each of which with 10^6^ iterations, starting from the posterior mode. To find the posterior mode we used the BFGS constrained minimisation algorithm, applied to − ln(*ℒ*) − ln(π), via the Matlab ^®^ function *fmincon*. In order to find a global minimum, we applied this method starting from 4000 random initial values.

## 3 Results

### Model fit

Denote by 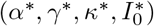 the posterior mode, and *S*\*(*t*), *I*\*(*t*), *R*\*(*t*), *D*\*(*t*) the solutions of the system (1) associated with these parameter values. The observation model (2) implies that the associated expected number of cases tested positive on day *t* is 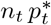 (expectation of a binomial) with

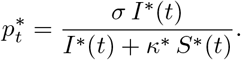

The observation model (3) implies that the expected cumulated number of deaths on day *t* is *D*\*(*t*).

To assess model fit, we compared these expectations and the observations, i.e., the cumulated number of cases tested positive,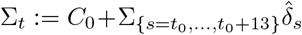 with *C*_0_ the number of cases tested positive by March 31 (*C*_0_ = 52 128) and the cumulated number of deaths 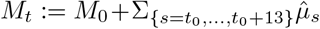, with *M*_0_ the number of reported deaths (at hospital) by March 31 (*M*_0_ = 3 123). The results are presented in Fig. 1. We observe a good match with the data.

**Figure 1:**
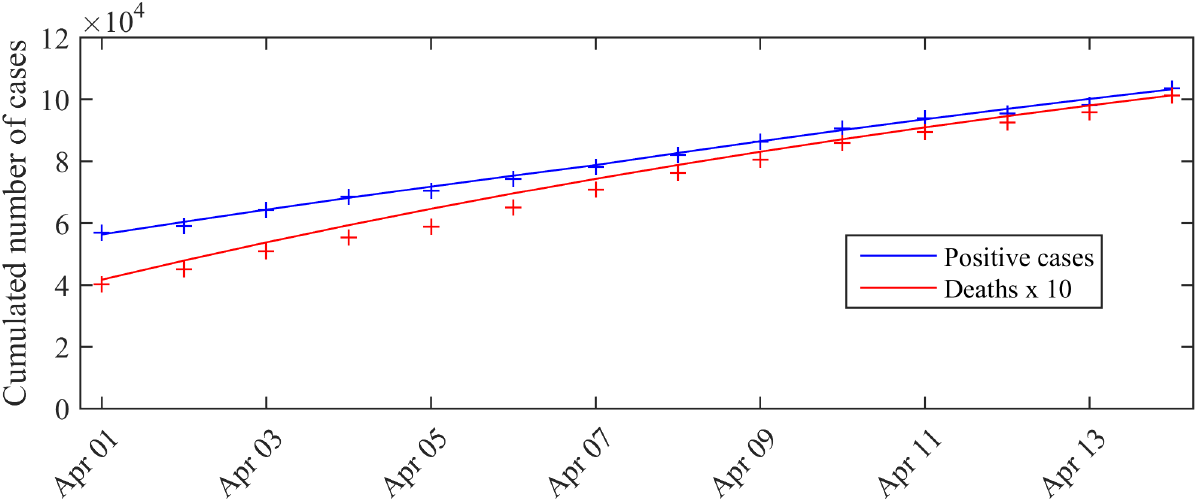
Expected number of observed cases and deaths associated with the posterior mode *vs* number of cases actually detected (total cases). The blue curve corresponds to the expected number of cases tested positive 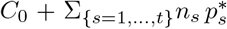 given by the model, the red curve corresponds to the expected cumulated number of deaths *D*\*(*t*) (excluding nursing homes). The crosses correspond to the observations (blue crosses: cumulated number of positive cases, red crosses: cumulated number of deaths). *C*_0_ is the number of cases tested positive on March 31 (*C*_0_ = 52 128).

The pairwise posterior distributions of the parameters (*α, I*_0_), (*α, γ*), (*α, κ*), (*γ, I*_0_), (*γ, κ*), (*κ, I*_0_) are depicted in Appendix, Fig. S2. With the exception of the parameter *γ* (Fig. S1), for which we chose an informative prior, the posterior distribution is clearly different from the prior distribution, showing that new information was indeed contained in the data.

### Contact rate and effective reproduction number

The effective reproduction number can be simply derived from the relation *R*_*e*_ = *α/β* [3]. The distribution of *R*_*e*_ is therefore easily derived from the marginal posterior distribution of the contact rate *α* (since we assumed *β* = 1*/*10; see Section 2.2). It is depicted in Fig. 2. We observe a mean value of *R*_*e*_ of 0.47 (95%-CI: 0.45-0.50).

**Figure 2:**
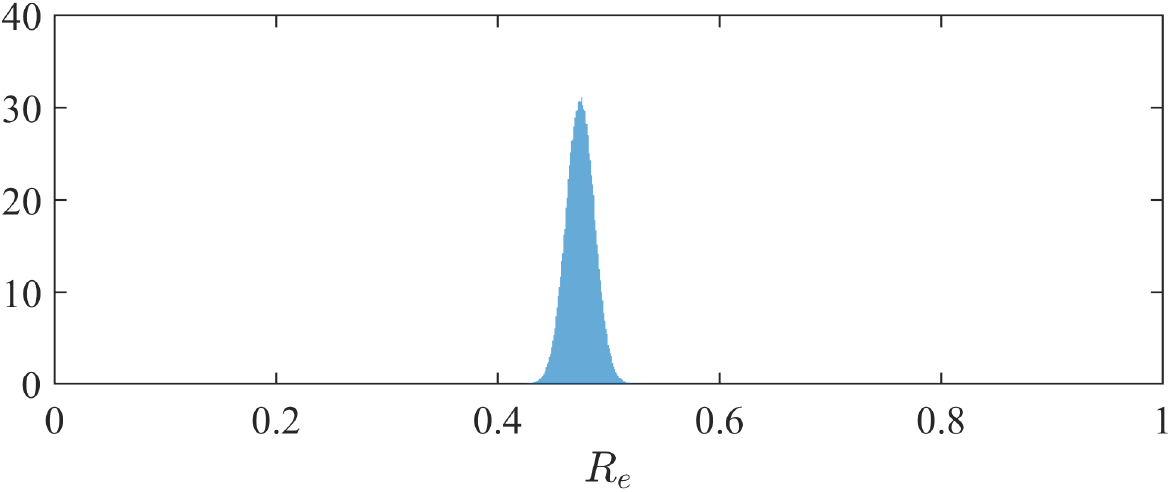
Posterior distribution of the effective reproduction number *R*_*e*_ in France.

### Dynamics of the infectious class

The marginal posterior distribution of *I*_0_ indicates that the number of infectious individuals at the beginning of the considered period (i.e. April 1st) is 1.4 *·* 10^6^ (95%-CI: 1.1 *·* 10^6^ − 1.8 *·* 10^6^). The computation of the solution of (1) with the posterior distribution of the parameters leads to a number of infectious *I*(*t*_*f*_) = 7.0 *·* 10^5^ and a total number of infected cases (including recovered) (*I* +*R*)(*t*_*f*_) = 2.0*·*10^6^ at the end of the observation period (April 14). By May 10, if the restriction policies remain unchanged, we get a forecast of *I*(*T*) = 1.6 *·* 10^5^ infectious cases (95%-CI: 1.3 *·* 10^5^ − 2.1 *·* 10^5^) and (*I* + *R*)(*T*) = 2.5 *·* 10^6^ infected cases including recovered (95%-CI: 2.0 *·* 10^6^ − 3.2 *·* 10^6^). The dynamics of the distributions of *I* and *I* + *R* are depicted in

Fig. 3. By May 10, the total number of infected cases (including recovered) therefore corresponds to a fraction of 3.7% of the total French population. This value does not include the recovered cases before April 1st.

**Figure 3:**
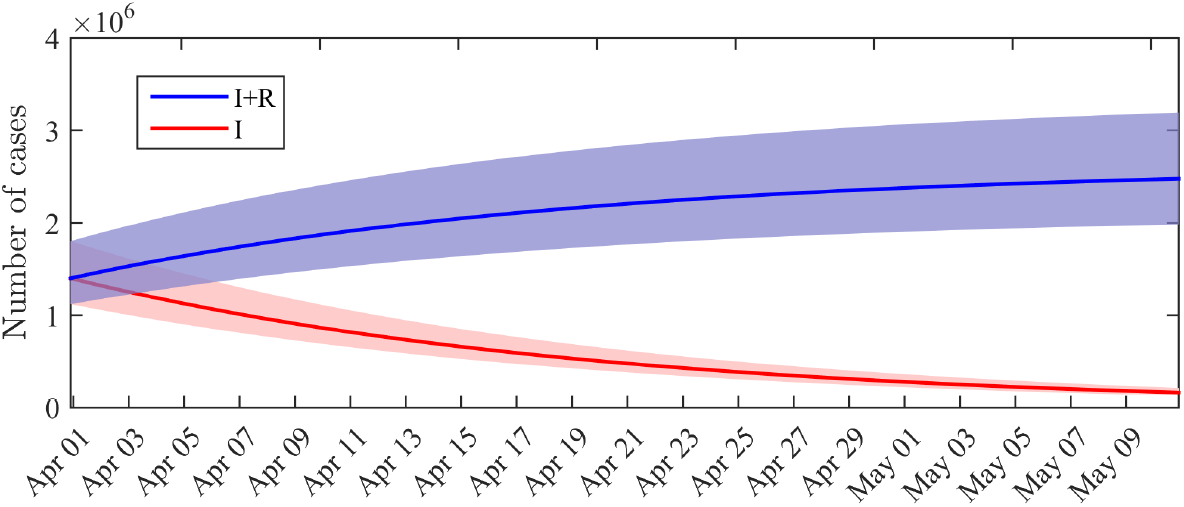
Distribution of the number of infectious cases *I*(*t*) and cumulated number of infected cases *I*(*t*) + *R*(*t*) across time. Solid lines: average value obtained from the posterior distribution of the parameters. Shaded areas: 0.025-0.975 interquantile ranges.

## 4 Discussion

Many studies focused on the estimation of the basic reproductive number *R*_0_ of the COVID-19 epidemic, based on data-driven methods and mathematical models (e.g., [21, 4]) describing the epidemic from its beginning. In average, the estimated value of *R*_0_ was about 3.3. We focused here on an observation period that began after the lockdown was set in France.

We obtained an effective reproduction number that was divided by a factor 7, compared to the estimate of the *R*_0_ carried out in France at the early stage of the epidemic, before the country went into lockdown (a value *R*_0_ = 3.2 was obtained in [12]). This indicates that the restriction policies were very efficient in decreasing the contact rate and therefore the number of infectious cases. In particular, the value *R*_*e*_ = 0.47 is significantly below the threshold value 1 were the epidemic starts dying out.

The decay in the number of infectious cases can also be observed from our simulations. It has to be noted that, although the number of infectious cases is a latent, or ‘unobserved’ process, the mechanistic-statistical framework allowed us to estimate its value (Fig. 3). The cumulated number of infected cases that we obtained by May 10 (*I* + *R*) corresponds to a fraction of 3.7% (95%-CI: 3.0-4.8%) of the total French population, without taking into account the number of recovered individuals before April 1st, which is not known. Based on a value *R*_0_ = 3.2, the herd immunity threshold, corresponding to the minimum fraction of the population that must have immunity to stop the epidemic, would be 1 − 1*/R*_0_ ≈ 69% (a threshold of 80% was proposed in [22]). This proportion will probably not be reached by May 10. As emphasised by [23], a too fast relaxation of the lockdown-related restrictions before herd immunity is reached or efficient prophylaxis is developed), would expose the population to an uncontrolled second wave of infection. In the worst-case scenario, the effective reproduction number *R*_*e*_ would approach the initially estimated value of *R*_0_, and the second wave would start with about 1.6 *·* 10^5^ infectious individuals (in comparison with the few cases that initiated the first wave in France) and about 64 *·* 10^6^ susceptible individuals. Keeping a low value of *R*_*e*_ is therefore crucial to avoid the saturation of hospital facilities.

## Data Availability

All data in the manuscript are available from opebn sources

## Author Contributions

L.R., E.K.K., J.P., A.S. and S.S. conceived the model and designed the statistical analysis. L.R. and S.S. wrote the paper, L.R. carried out the numerical computations. All authors reviewed the manuscript.

---

**Figure S1:**
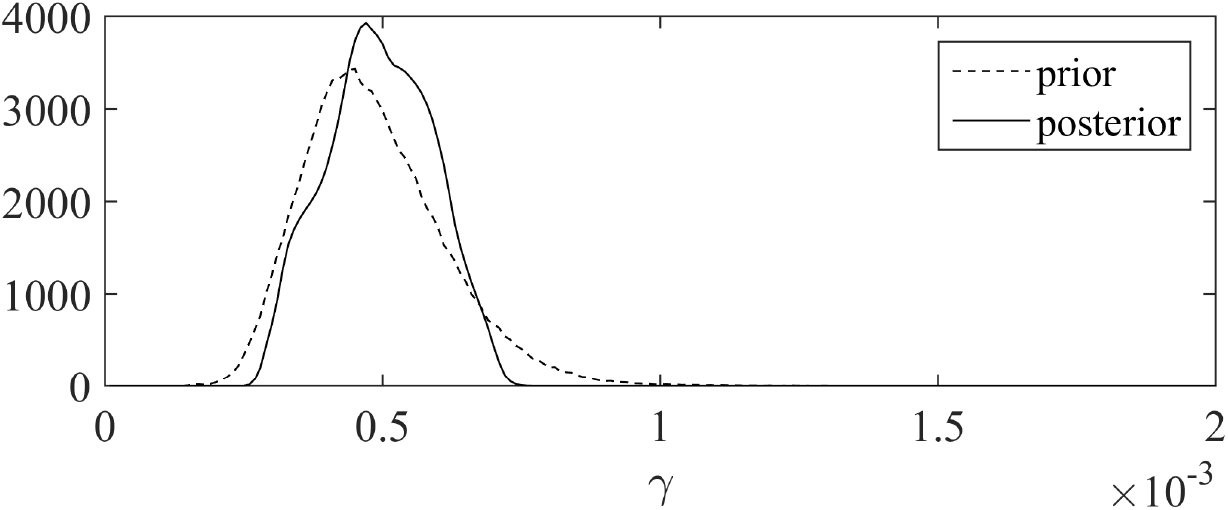
Prior and marginal posterior distributions of the death rate *γ*.

## Appendix

- The prior and marginal posterior distributions of the death rate *γ* are depicted in Fig. S1.
- The pairwise posterior distributions of the parameters (*α, I*_0_), (*α, γ*), (*α, κ*), (*γ, I*_0_), (*γ, κ*), (*κ, I*_0_) are depicted in Fig. S2.

**Figure S2:**
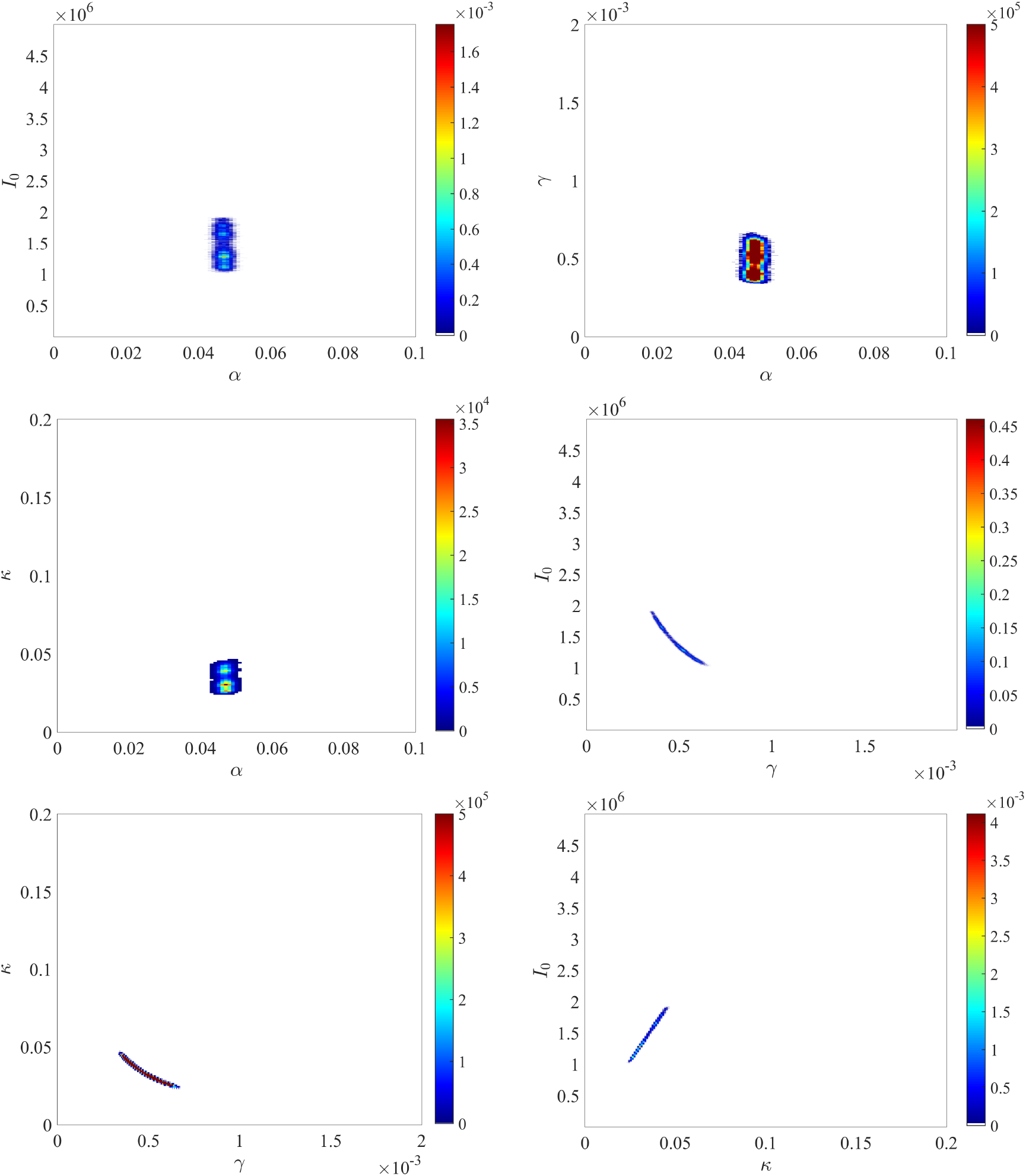
Joint posterior distributions of (*α, I*_0_), (*α, γ*), (*α, κ*), (*γ, I*_0_), (*γ, κ*) and (*κ, I*_0_).

